# The global burden of HIV among Long-distance truck drivers: A systematic review and meta-analysis

**DOI:** 10.1101/2023.12.18.23300177

**Authors:** Cyrus Mutie, Berrick Otieno, Elijah Mwangi, Rosemary Kawira, Albanus Mutisya, John Gachohi, Grace Mbuthia

## Abstract

Long-distance truck drivers (LDTDs) endure a disproportionately high burden of HIV in various global settings. However, unlike other most at-risk populations, the global burden of HIV among LDTDs has not been documented so far. The result has been poor allocation and distribution of the limited HIV preventive resources for LDTDs in most parts of the world. Thus, a systematic review and meta-analysis were conducted to assess the global burden of HIV among LDTDs. A comprehensive electronic search was conducted in PubMed, ProQuest Central, PubMed Central, CINAHL, and Global Index Medicus to identify relevant information published in English on HIV prevalence among LDTDs from 1989 to the 16^th^ of May 2023. A random-effects meta-analysis was conducted to establish the burden of HIV at global and regional levels. The Joanna Briggs Institute (JBI) and Newcastle-Ottawa Scale (NOS) tools were used to assess the quality of the included studies.

Of the 1787 articles identified, 43 were included. Most of the included studies were conducted in sub-Saharan Africa (44.19%, n=19), and Asia and the Pacific (37.21%, n=16). The pooled prevalence of HIV was 3.82%. The burden of HIV was highest in sub-Saharan Africa at 14.34%, followed by Asia and the Pacific at 2.14%, and lastly Western, Central Europe and North America at 0.17%. The overall heterogeneity score was (*I*^2^ = 98.2%, p < 0.001).

The global burden of HIV among LDTDs is 3.82%, six times higher than that of the general population globally. Compared to other regions, the burden of HIV is highest in sub-Saharan Africa at 14.34%, where it’s estimated to be 3% in the general population. Thus, LDTDs endure a disproportionately high burden of HIV compared to other populations. Consequently, more LDTD-centred HIV research and surveillance is needed at national and regional levels to institute tailored preventive policies and interventions.

**PROSPERO Number:** CRD42023429390

## Introduction

Globally, the incidence rate of HIV has declined by 38%, from 2.1 million in 2010 to 1.3 million in 2022 [1]. While significant declines as high as 57% have occurred in Eastern and Southern Africa, the incidence rate of HIV has increased by 61% in the Middle East and North Africa, and 49% in Eastern Europe and Central Asia [1]. The declines point to key milestones that have so far been made in the control and prevention of HIV/AIDS, however, the increasing incidence rates in some parts of the world may derail the global efforts to end the epidemic [1]. Moreover, the declines may mask the disproportionately high burden of HIV endured among certain key population groups like female sex workers (FSWs) [2] and the hard-to-reach populations like the Long-distance truck drivers (LDTDs) [3,4].

Studies have identified several factors likely to drive a high burden of HIV among LDTDs. These include; sexual risk behaviours like poor condom use [5–8], multiple sexual partners [9–11], illicit drug and alcohol use during or before sexual interactions [10,12–14], and risky sexual networks [15–18]. Notably, LDTDs’ stopovers along their transit routes are characterized by prevalent commercial sex from FSWs whose risk of HIV infection is also high [19]. Indeed, according to UNAIDS, FSWs were thirty times more likely to contract HIV in 2021, positioning their sexual clients like LDTDs at a high-risk profile of contracting and spreading the infection [20]. The LDTDs’ stopovers often lack proper HIV prevention measures, such as access to condoms, information, and HIV testing services, further exacerbating the risk of transmission [21]. Additionally, mental exhaustion and psychosocial problems due to long working hours while in transit have been implicated as predictors of risky sexual behaviours likely to elevate the burden of HIV among LDTDs [22–25]. Moreover, LDTDs’ varying eligibility for HIV preventive services across different geographic borders [26], constant mobility [27,28], and missed clinic appointments [21,24,29] limit their access to essential preventive services, further predisposing them to the risk of HIV.

The burden of HIV among LDTDs has far-reaching health implications. In addition to the negative health outcomes for the drivers themselves, their frequent contact with other individuals, including their partners, FSWs, and other populations along transit routes, increases the risk of HIV transmission to the general population. Unlike other high-risk groups like FSWs, there has been paucity of information on the burden of HIV among the LDTDs at global, regional and national levels.

The undetermined burden of HIV among LDTDs is most likely to be succeeded by a lack of targeted HIV prevention interventions, resulting in poor use and allocation of scarce health resources. Hitherto, no systematic review has been done to quantify the global burden of HIV among LDTDs. Previous reviews have summarized the prevalence of HIV among LDTDs at national levels in Ethiopia [30] and China [31], limiting the generalizability of their findings to other global settings. Therefore, with the increasing efforts to fight HIV, especially among the most at-risk populations, an extensive review is warranted to put into perspective the global burden of HIV among LDTDs. To address the aforementioned gap, this study set out to systematically summarize and analyse the available evidence on HIV prevalence among LDTDs from various parts of the world.

A review of this kind may amplify the evidence needed to guide surveillance, and allocation of preventive resources, strategies and policies to reduce the burden of HIV among LDTDs in various parts of the world. More specifically, this review contributes to achieving the UNAIDS 95-95-95 targets by the year 2025 and the global goal to end AIDS by 2030 [1,20].

## Methods

### Protocol and Registration

A protocol for this study was developed and registered in the International Prospective Register of Systematic Reviews (PROSPERO) under registration number CRD42023429390. The study followed the Preferred Reporting Items for Systematic Review and Meta-analysis (PRISMA) guidelines [32]. A PRISMA checklist is provided (S1 Table)

### Eligibility Criteria

Articles identified from the electronic search had to meet pre-determined inclusion and exclusion criteria to be included. The inclusion and exclusion criteria were categorized into condition (HIV), context (where the studies were conducted), population of interest (Long-distance truck drivers) and the study design. A summary of the inclusion and exclusion criteria for the selected studies is given in Table 1.

**Table 1.** Inclusion and exclusion criteria for selected studies.

| Inclusion | Exclusion |
| --- | --- |
| **Condition**<br>Studies on prevalence, frequency, proportion, burden or risk of HIV.<br>Studies reporting on sexually transmitted infections, HIV included.<br>Studies reporting on the specific validated HIV laboratory testing method. | Studies not specific on the quantification of HIV (not specific on prevalence, frequency, proportion, burden or risk of HIV).<br>Studies reporting prevalence of other sexually transmitted infections without inclusion of HIV. |
| **Context**<br>Studies that are specific on the region/country/place in which the study was conducted. | Studies not specific on which parts of the world the data was collected. |
| **Population**<br>Studies reporting HIV status among long-distance truck drivers from all parts of the world.<br>Studies reporting on HIV status of assistants of long-distance truck drivers. | Studies reporting HIV status among drivers not covering long distances.<br>Studies reporting HIV status among drivers of other vehicles besides long distance trucks.<br>Studies only reporting HIV status of sexual partners of long-distance truck drivers.<br>Studies reporting on HIV- status of assistants of long-distance truck drivers not covering long distances. |
| **Studies**<br>Cross-sectional studies.<br>Studies with specified sample sizes.<br>Cohort studies.<br>Interventional studies (baseline outcomes). | Review papers (scoping reviews, systemic reviews and meta-analysis).<br>Case-reports.<br>Case-series studies.<br>Studies with unspecified sample sizes.<br>Books and Book reviews.<br>Qualitative studies (phenomenological studies, ethnographic studies, grounded theories).<br>Publications from conference proceedings, workshops and seminars. |

### Information sources

A comprehensive structured electronic search was conducted in PubMed, PMC (PubMed Central), CINAHL (Cumulated Index to Nursing and Allied Health Literature), ProQuest Central, and Global Index Medicus to identify all relevant studies published since inception to 16^th^ of May 2023 (when the last search was conducted). Additional studies were manually searched from the list of references available from the full-text articles retrieved.

### Search strategy

The basic search terms were divided into three categories: 1. Condition (HIV OR AIDS OR HIV/AIDS OR Sexually transmitted diseases; 2. Population (Long-distance truck drivers OR Long-distance truckers OR Long-haul truck drivers; and 3. Outcome measures (prevalence OR burden OR risk). Synonyms in each of the search categories were combined with the Boolean operator “OR”. The three search categories were combined with the Boolean operator “AND” to form the following search string (HIV OR AIDS OR HIV/AIDS OR “Human immunodeficiency virus” OR “Immune suppression” OR “Immuno-suppression” OR “Acquired immune deficiency syndrome” OR “sexually transmitted diseases” OR “sexually transmitted infections” OR “venereal diseases” OR “venereal infections”) AND (prevalence OR burden OR proportion OR epidemiology OR frequency OR risk) AND (Truckers OR “Long-distance truckers” OR “Long-distance truck drivers” OR “Long-haul drivers” OR “Long-haul truck drivers” OR “Long-haul truck assistants” OR “Long-distance truck assistants” OR “Migrant truck drivers” OR “Migrant truck assistants”). Medical Subject Headings (MeSH) terms were also used where applicable. Where the search was unsuitable for a database, it was modified accordingly. All searches were restricted to studies published in English. Search strings from PubMed and PMC databases are provided (S1 File).

### Selection process

Two reviewers C.M, and B.O. independently screened the records for eligibility in three stages. In the first stage, deduplication was done using the Zotero bibliography software. This was followed by the screening of titles and abstracts using the Rayyan online platform [33]. In the third stage, full-text articles were assessed for eligibility. Illegible studies were removed, and their bibliographic details listed with specific reasons. Any disagreements between the two reviewers were resolved by a third author G.M.

### Data extraction and data items

Using a standardized Microsoft Excel data extraction form, C.M independently extracted the following variables from the retrieved articles: (a) details of the retrieved articles (author and year of publication); (b) details of the study (year the study was conducted, country, continent, region, economic class of the country of origin, community or hospital-based, and study design); (c) characteristics of study participants (sample size, gender, and age); (d) measures of outcome (laboratory method used to diagnose HIV, number of HIV positives, HIV prevalence, levels of HIV infection compared to the general population, predicting factors and quality scores). Thereafter, B.O. went through the extracted data, flagged possible anomalies and resolved them through a consensus with C.M. These variables were coded appropriately in an Excel spreadsheet, to enable ease in data synthesis and analysis.

### Risk of bias and quality assessment

Two reviewers, C.M. and B.O. independently assessed the risk of bias and quality for each study. This was done using the Joanna Briggs Institute (JBI) critical appraisal tools [34,35] for cross-sectional (prevalence) studies and non-randomized interventional (quasi-experimental) studies, and the Newcastle-Ottawa Scale (NOS) for assessing the quality and bias of cohort studies [36]. The quality of the prevalence studies was graded based on nine items in the JBI checklist. Following the NOS recommendations, the grading of the cohort studies was based on stars and categorized as follows; 0-4 Unsatisfactory; 5-6 Satisfactory; 7-8 Good; and 9-10 Very good. Additionally, publication bias was assessed using funnel plots (Fig 3).

### Strategy for data synthesis

The reviewers derived individual study proportions by dividing the number of HIV-positive cases reported by the size of the sample studied and then computed the overall crude prevalence estimates expressed per 100 participants, for the positive cases stratified according to age and study region among other potential correlates. To obtain a more precise estimate of the overall prevalence of HIV and to assess for heterogeneity across studies, a meta-analysis of prevalences using the random-effects model was performed using R version 4.3.1 [37]. The researchers transformed reported prevalences using logit transformation to have them follow a normal distribution, hence ensuring an accurate estimate of the summary prevalences, where the logit transformation of the prevalence and its inverse variance weight were calculated as; *ln* (*p*/1-*p*) and *wl* = *np* (1-p) respectively [38].

After the analysis, the transformed prevalence and the confidence intervals were back-transformed to prevalence for reporting. A formal χ2 test with a Q-statistic was used to assess for study heterogeneity under a null hypothesis of homogeneity among the study effect sizes [39], as heterogeneity could result from study sampling error or between studies’ variability due to true effect size differences across reported studies. Also, because of the variation in methodological approaches, sample sizes, and the study settings, heterogeneity was anticipated. The researchers applied a random-effects model to quantify the degree of heterogeneity using the *I^2^* statistic, where it was assumed that *I^2^* values of 25, 50, and 75% indicate low, moderate, and high heterogeneity, respectively, as the values of *I^2^* increase with increasing levels of heterogeneity [40]. Forest plot was used to visualize the heterogeneity among studies, prevalence estimates, and their confidence intervals. Visual inspection of the degree of asymmetry was done using funnel plots to assess potential publication bias. Further, a univariable and multivariable meta-regression analyses were done to investigate factors attributable to heterogeneity.

### Analysis of subgroups or subsets

The reviewers investigated potential factors that could explain the variability in prevalence estimates across the studies when *I^2^* values are substantial using meta-regression univariable and multivariable models. This was performed using logit transformed proportions, individual effect sizes, and their corresponding sampling variances. Factors such as year of study publication, the continent of the study, country income classification, study setting, age group, country of study, measurement tool used, , sample size, and quality score of the study were investigated to examine and quantify the magnitude of their impact on prevalence estimates.

## Results

### Study Selection

The initial electronic search yielded a total of 1787 articles from five databases (PubMed, n=150; PMC, n=746; ProQuest Central, n=834; CINAHL, n=51, and Global Index Medicus, n=6). An additional 12 articles were identified from the reference list and Google Scholar. A total of 63 duplicates were removed, after which 1736 articles were screened for eligibility. Out of these, 1634 irrelevant records were excluded based on their title and abstract. After screening the remaining 102 full texts, 43 studies were included in the review and meta-analysis. A PRISMA flow chart for the systematic review process is shown in Fig 1.

**Fig 1.**
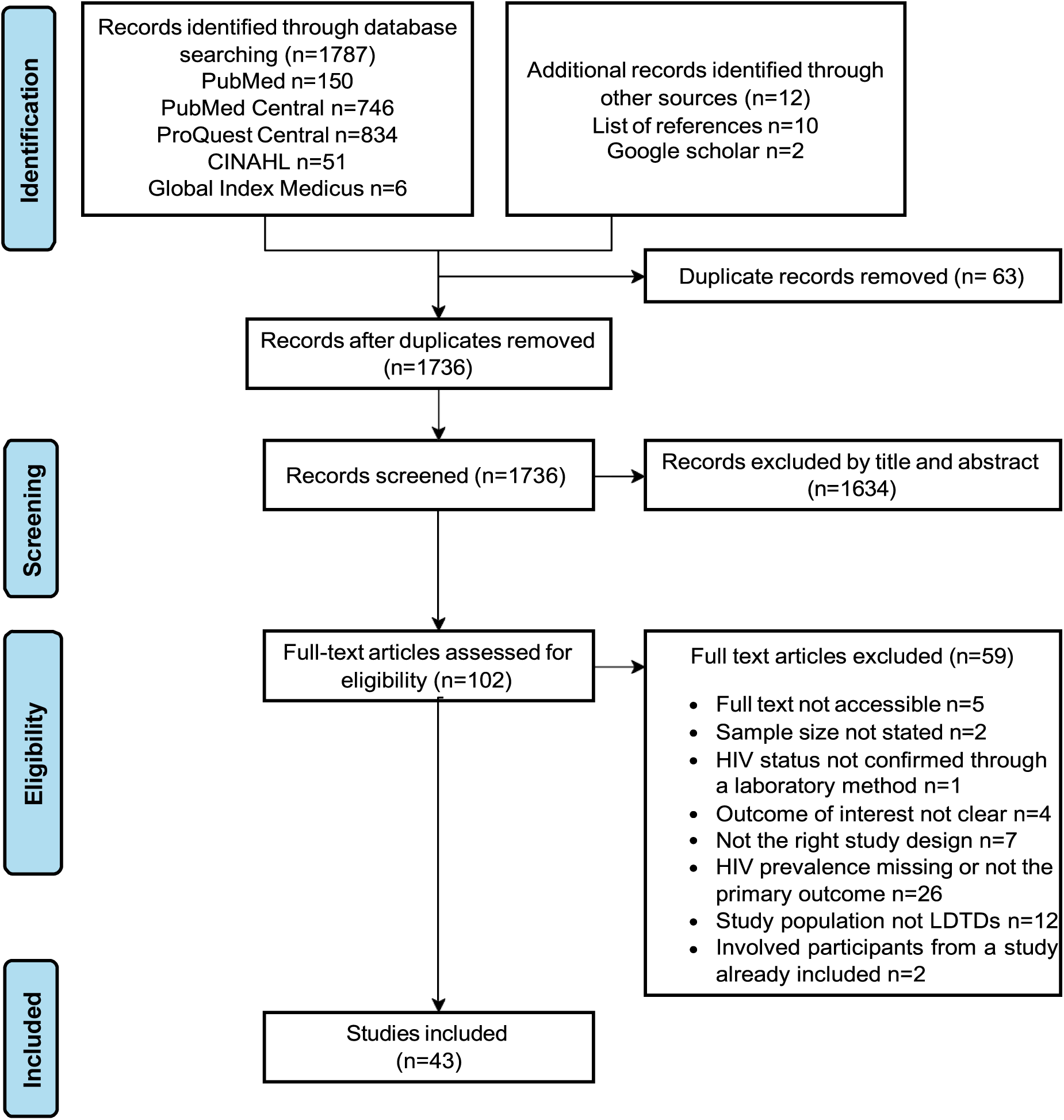
PRISMA flowchart illustrating the systematic review process.

### Study characteristics

A detailed summary of the 43 included studies is provided (S2 Table). Most of the included studies (65.12%, n=28), were published between 1989 and 2010, except for (34.81%, n=15) published from 2011 and onwards. A majority of the included studies were conducted in sub-Saharan Africa (44.19%, n=19), followed by Asia and the Pacific (37.21%, n=16), with Eastern Europe and Central Asia trailing behind with only one study. Studies from sub-Saharan Africa were mostly conducted in Kenya (n=5), and South Africa (n=4), whereas in Asia and the Pacific, most of them were conducted in India (n=12). A map showing the geographical distribution of the studies is given in Fig 2. Almost all of the included studies (90.7%, n=39) were cross-sectional by design [3,9,11,27,41–75] except for two prospective cohort studies [76,77] and two non-randomized interventional studies [78,79].

**Fig 2.**
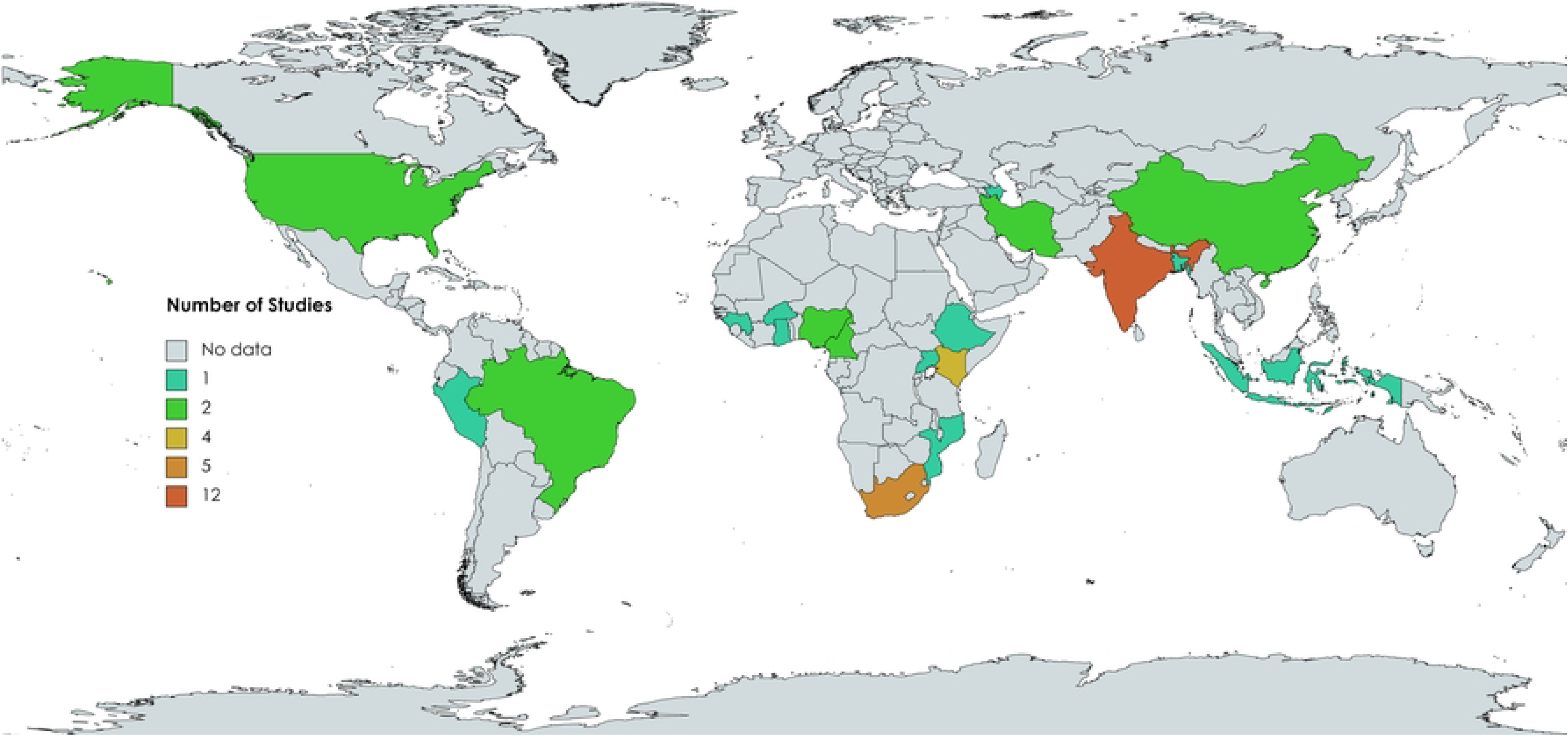
Global distribution of the included studies for prevalence of HIV among long-distance truck drivers

**Fig 3.**
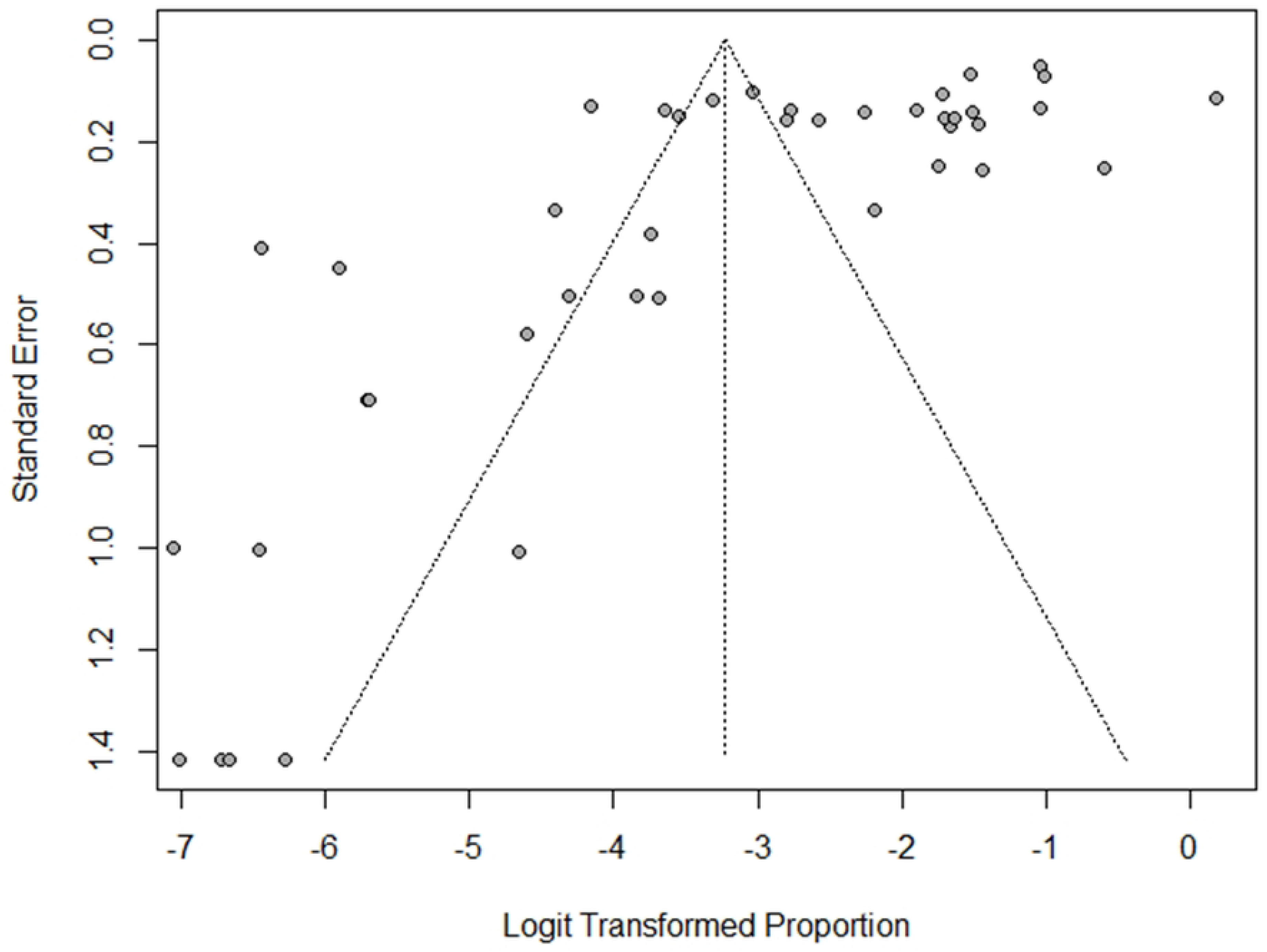
Funnel plot showing precision of the 43 included studies on global burden of HIV among Long-distance truck drivers

### Study population

The 43 included studies comprised a total sample size of 34,847, ranging from 68 [50] to 3,763 [47] by individual studies. By region, the highest sample size was Asia and the Pacific (n=15,871, 45.54%) followed by sub-Saharan Africa (n=10,014, 28.74%). Only one included study [75] involved both male and female LDTDs, whereas the rest had predominantly male participants. Most of the included studies (79.07%, n=34) targeted LDTDs as the primary study population, except for [45,58,59,63,66–68,76,77], which involved a mixture of LDTDs and other study populations (S2 Table).

### Laboratory method of HIV diagnosis

The enzyme-linked immune-sorbent assay (ELISA) was widely used as the laboratory method of diagnosing HIV in most (74.42%, n=32) of the included studies, except for [41,50,51,54,66,73,75] where a combination of ELISA and Western Blot Assay technologies were used. Other individual studies used enzyme immunoassay (EIA) and Innotest HIV-1/HIV-2 [64], Orasure [3], Determine & Oraquick [47], and OraQuick [44] (S2 Table).

### HIV Prevalence estimates

The pooled prevalence of HIV among LDTDs in all global regions was 3.82% (95% CI 2.22-6.49) as shown in table 2. In the sub-group analysis by region, the highest prevalence of HIV among LDTDs was from sub-Saharan Africa 14.34% (95% CI 9.94-20.26), followed by Asia and Pacific 2.14% (95% CI 1.01-4.51), Eastern Europe and Central Asia 1.54% (95% CI 1.17-1.99), Latin America and Caribbean 0.37% (95% CI 0.08-1.60), Middle East and North Africa 0.27% (95% CI 0.08-0.93), and lastly Western, Central Europe and North America registering the lowest estimates of 0.17% (95% CI -0.03-0.82). In sub-Saharan Africa highest prevalence rates (above 15%) were reported in South Africa [3,72], Uganda [50], Kenya [49,65,77], Burkina Faso [62] and Cameroon [67]. In the Asia and Pacific region, the highest HIV prevalence estimates were from India [11,55,58,64,69], whereas China registered the lowest prevalence rates [51]. In Latin America and the Caribbean, the highest HIV prevalence estimates were from Brazil [60]. In the other regions, there were no wide variations in the HIV prevalence reported from individual countries. Details of the pooled prevalence in the different studies are given in Table 2. Forest plots showing more details of sub-group analysis are given (S3 Table).

**Table 2.** Pooled prevalence and subgroup analysis of the burden of HIV among long-distance truck drivers.

| Classification | Categories | Number of Studies<br>n (%) | Number of HIV Infections<br>(n) | Prevalence of HIV<br>(%) | 95% CI | HIV Negative<br>n (%) | Total Sample<br>Size<br>n (%) | Heterogeneity (I<sup>2</sup>)<br>Score<br>%, p-value |
| --- | --- | --- | --- | --- | --- | --- | --- | --- |
| **Overall** |  | 43 | 2309 | 3.82 | 2.22-6.49 | 32538 (93.4) | 34847 | 98.2, p < 0.001 |
| **Continent** | Africa | 19 (44.19) | 1774 | 14.34 | 9.94-20.26 | 8240 (82.3) | 10014 (28.74) | 97.0, p < 0.01 |
|  | Asia | 19 (44.19) | 524 | 1.75 | 0.87-3.51 | 20124 (97.5) | 20648 (59.25) | 96.5, p < 0.01 |
|  | North America | 2 (4.65) | 1 | 0.17 | 0.03-0.82 | 901 (99.9) | 902 (2.59) | 0.0, p = 0.92 |
|  | South America | 3 (6.98) | 10 | 0.37 | 0.8-1.60 | 3273 (99.7) | 3283 (9.42) | 77.0, p = 0.01 |
| **UNAIDS Region** | Asia and the Pacific | 16 (37.21) | 464 | 2.14 | 1.01-4.51 | 15407 (97.08) | 15871 (45.54) | 96, p < 0.01 |
|  | Eastern Europe and Central Asia | 1 (2.33) | 58 | 1.54 | 1.17-1.99 | 3705 (98.46) | 3763 (10.8) | NA |
|  | Latin America and The Caribbean | 3 (6.98) | 10 | 0.37 | 0.08-1.60 | 3273 (96.7) | 3283 (9.42) | 77, p = 0.01 |
|  | Middle East and North Africa | 2 (4.65) | 2 | 0.27 | 0.08-0.93 | 1012 (99.80) | 1014 (2.91) | 0, p = 0.52 |
|  | Sub-Saharan Africa | 19 (44.19) | 1774 | 14.34 | 9.94-20.26 | 8240 (82.3) | 10014 (28.74) | 97.0, p < 0.01 |
|  | Western, Central Europe and North America | 2 (4.65) | 1 | 0.17 | 0.03-0.82 | 901 (99.89) | 902 (2.59) | 0%, p = 0.92 |

### Heterogeneity of Included Studies

The overall heterogeneity score was (*I*^2^ = 98.2, p < 0.001) indicating that the 43 studies included were highly heterogeneous. By regions, the highest heterogeneity was from studies conducted in Sub-Saharan Africa (*I*^2^= 97.0, p < 0.01), followed by those from Asia and the Pacific (*I^2^*= 96, p < 0.01). Studies from the Middle East and North Africa, Eastern Europe and Central Asia and Western, Central Europe and North America had the lowest level of heterogeneity though not statistically significant as shown in Table 3 and Fig 3. Further investigation revealed that; the year of publication (aβ -0.047; 95% CI (-0.089; -0.005); p=0.027) a study being conducted in the regions of sub-Saharan Africa (aβ 1.719; 95% CI (0.939; 2.50); p<.0001), Latin America and the Caribbean (aβ -1.89; 95% CI (-3.419; -0.360); p=0.016 and Western, Central Europe and North America (aβ -2.432; 95% CI (-4.732; -0.131); p=0.038, significantly accounted for heterogeneity in the included studies as shown in Table 3.

**Table 3.** Meta-regression analysis for factors attributed to heterogeneity in the 43 included studies.

| Factors | Univariable Analysis |  | Multivariable Analysis |  |
| --- | --- | --- | --- | --- |
| | Beta Coefficient [$\beta$ (95% CI)] | P-Value | Adjusted Beta Coefficient [$a\beta$ (95% CI)] | P-Value |
| Year of Publication | **-0.0926 (-0.1496; -0.0356 )** | **0.0015** | **-0.0471 (-0.0889; -0.00530)** | **0.0272** |
| Cross-sectional | 0 (Reference) |  | (Reference) |  |
| Integrated Biological and Behavioral Surveillance Survey | 1.3054 (-2.3415; 4.9524) | 0.4829 | - | - |
| Interventional Study (Baseline) | -0.8257 (-4.5925; 2.9410) | 0.6674 | - | - |
| National Cross-sectional Survey | -0.7883 (-2.4453; 0.8687) | 0.3511 | - | - |
| National Sentinel Survey | -1.3328 (-3.5352; 0.8696) | 0.2356 | - | - |
| Prospective-Cohort (after follow up) | 0.2142 (-3.4331; 3.8614) | 0.9084 | - | - |
| Prospective-Cohort (baseline) | 1.4784 (-2.1583; 5.1150) | 0.4256 | - | - |
| Quasi-experimental (Baseline) | -2.8932 (-6.6319; 0.8455) | 0.1293 | - | - |
| Africa | (Reference) |  | (Reference) |  |
| Asia | **-2.1298 (-2.9251; -1.3346)** | **<.0001** | - | - |
| North-America | **-4.5640 (-6.9469; -2.1810)** | **0.0002** | - | - |
| South-America | **-3.7862 (-5.4022; -2.1702)** | **<.0001** | - | - |
| Asia and the Pacific | (Reference) |  | (Reference) |  |
| Eastern Europe and Central Asia | -0.4110 (-2.7547; 1.9326) | 0.7310 | -0.3783 (-2.5765; 1.8198) | 0.7359 |
| Latin America and The Caribbean | **-1.8525 (-3.4633; -0.2418)** | **0.0242** | **-1.8894 (-3.4190; -0.3599)** | **0.0155** |
| Middle East and North Africa | **-2.3156 (-4.5252; -0.1059)** | **0.0400** | -1.8859 (-4.0435; 0.2717) | 0.0867 |
| Sub-Saharan Africa | **1.9314 (1.1236; 2.7391)** | **<.0001** | **1.7193 (0.9389; 2.4998)** | **<.0001** |
| Western, Central Europe and North America | **-2.6357 (-5.0051; -0.2663)** | **0.0292** | **-2.4319 (-4.7323; -0.1316)** | **0.0383** |
| Mean Age | -0.0904 (-0.2745; 0.0937 ) | 0.3358 | - | - |
| Sample Size | -0.0005 (-0.0011; 0.0001) | 0.0789 | - | - |
| Determine & Oraquick | (Reference) |  | (Reference) |  |
| EIA; Innostest HIV-1/HIV-2 | 2.4965 (-2.3420; 7.3350) | 0.3119 | - | - |
| ELISA | 0.9683 (-2.5096; 4.4463) | 0.5853 | - | - |
| ELISA & Western Blot Assay | -0.1153 (-3.8185; 3.5878) | 0.9513 | - | - |
| GAL ELISA HIV I/II | 4.3251 (-0.5073; 9.1575) | 0.0794 | - | - |
| OraQuick | -2.1215 (-7.6892; 3.4462 ) | 0.4552 | - | - |
| Orasure | 3.1110 (-1.7173; 7.9393) | 0.2066 | - | - |
| Rapid Assay Test | 1.8902 (-2.9451; 6.7256) | 0.4436 | - | - |
| Quality Score | -0.2063 (-0.5741; 0.1616) | 0.2718 | - | - |

### Bias and Quality Scores of the Included Studies

Of the 39 cross-sectional studies included, 11 were of high quality (low risk of bias) [11,27,49,51,56,64,68–72], extra 11 were of good quality (low risk of bias) [3,45–47,52–54,59,61,73,75], 4 were of low quality (high risk of bias) [9,62,65], whereas the remaining 17 had a fair rating (moderate risk of bias). All the cohort [76,77] and interventional [78,79] studies included were of high-quality rating (low risk of bias). Three tables for the quality scores of the included cross-sectional, cohort and non-randomized interventional studies were generated (S4-S6 Tables). The generated funnel plot was asymmetrical implying the presence of publication bias in the included studies as indicated in Fig 3.

## Discussion

This systematic review and meta-analysis purposed to quantify the global burden of HIV infections among LDTDs. The findings from this study demonstrate substantial evidence of high levels of HIV across LDTDs of diverse geographical backgrounds such as sub-Saharan Africa, Asia and the Pacific. While for a long time LDTDs were understood to be both most at-risk [5,6,9,13,15,17,19,80] and a hard-to-reach population [3,4,18,27], this is even the pioneer study to systematically determine and report the extent of the burden of HIV in this key population.

The pooled prevalence of HIV was 3.82% (95% CI 2.22-6.49). This is nearly six times the current global HIV prevalence of 0.7% in the adult general population based on the latest UNAIDS report [1]. This pooled prevalence is higher than those reported for other most-at-risk populations like sex workers (2.5%), and people in prisons (1.4%), but lower than that of people who inject drugs (5.0%), gay men and other men who have sex with men (MSMs) (7.7%), and transgender groups (10.3%) [1]. Moreover, the pooled prevalence was substantially higher compared to the prevalence of HIV in other migrant populations in high-income countries (2.25%) and low-income countries (0.23%) [81]. Nevertheless, a recent meta-analysis identified a prevalence of 17.3% among FSWs in the U.S. [82]. This finding demonstrated that LDTDs endure a high burden of HIV especially in sub-Saharan Africa, Asia and the Pacific regions. However, this assertion may not suffice for some regions like Latin America and the Caribbean, Eastern Europe and Central Asia, Middle East and North Africa and Western, Central Europe and North America, where a very low number of studies were identified. Indeed, the high burden of HIV may for long have been masked by the more attention given to other most-at-risk populations, somewhat isolating LDTDs in various HIV preventive and research programs owing to their hard-to-reach nature [83]. Furthermore, the high burden may infer delayed success of HIV prevention among this population, which consequentially derail the collective global efforts to fight the epidemic. Therefore, there is a need to scale up targeted interventions with an emphasis on combination prevention; a holistic approach with effective structural, biomedical and behavioural HIV prevention services among LDTDs as recommended by the UNAIDS [20].

In the current study, highest pooled prevalence of HIV among LTDs was in sub-Saharan Africa, followed by the Asia and Pacific region, and the least from Western, Central Europe and North America. This is so despite the sample size from sub-Saharan Africa (28.74%), being nearly a third that of the entire study. Thus, the high overall pooled prevalence observed is attributable to the sub-Saharan Africa region. This trend is consistent with the global burden of HIV in the general population which is highly concentrated in sub-Saharan Africa [1,20]. Indeed, the 14.34% HIV prevalence which is about five times that of the general population estimated to be around 3% in the region [1] mirror our global pooled prevalence which is almost six times that in general population. Moreover, a high burden of HIV among LDTDs was concentrated in the Eastern [48–50,65,77] and Southern [3,27,72] regions of sub-Saharan Africa, compared to the Western and Central Africa region, reflecting the UNAIDS report on HIV trends in the general population [1,20].

The high burden of HIV among LDTDs in sub-Saharan Africa may be attributed to various factors. First, studies have identified that LDTDs in sub-Saharan Africa mainly solicit commercial sex from FSWs, who equally share a disproportionately high burden of HIV [19,72,84]. Sexual interactions between LDTDs and commercial sex workers are known to be characterized by risky sexual behaviours [19], which increases the risk of HIV infection. Additionally, limited access and inadequate HIV preventive services among LDTDs along most transport routes in sub-Saharan Africa [3,61,85] , increase their vulnerability to HIV. Furthermore, due to limited healthcare resources in most sub-Saharan African countries, LDTDs may find themselves ineligible for HIV preventive services across different geographic borders [26]. The high burden of HIV among LDTDs in sub-Saharan Africa may partially be a driving factor to the high burden sustained in the region. Thus, in order to sustain the currently reducing HIV incidence rates in sub-Saharan Africa, enhanced targeted HIV preventive services among the LDTDs are needed to match those offered in other most-at-risk populations like the FSWs.

The 2.14% prevalence of HIV among LDTDs in Asia and the Pacific is above the estimates of below 1% in the general population in the region [1]. However, the prevalence reported by individual studies widely varied, with some studies from India reporting a prevalence of above 15% [55,58,64], compared to a prevalence of 0% in Bangladesh [56] and China [51]. Thus, this evidence also demonstrates the need for enhanced HIV preventive services among LDTDs in Asia and the Pacific.

There was a low number of studies identified from Latin America and the Caribbean, Western, Central Europe and North America, Middle East and North Africa and Eastern Europe and Central Asia, with each region having less than three studies. Ultimately, the data from these regions is inadequate to make a conclusive comparison in the context of the current study. This may be due to underreporting of HIV prevalence or a lack of studies that focus on HIV among LDTDs in those regions. Indeed, given the relative low burden of HIV in those regions [1], LDTDs’ centred research has been focused on other healthcare needs [12,44,86–89]. However, this school of thought may not be completely true, given the recent sharp increase in incidence rates of HIV in some of the regions mentioned above like the Middle East, North Africa and Eastern Europe [1]. Nevertheless, the HIV prevalence reported in the individual studies still demonstrates a substantial prevalence of HIV which needs intervention mechanisms. Thus, more LDTDs’ centred research and HIV surveillance activities are needed to guide the formulation of suitable targeted strategies, policies and interventions to prevent new infections among them.

This review identified an overall heterogeneity of 98.2%, p < 0.001 in the included studies. This was mainly attributable to the year of publication and studies being conducted in sub-Saharan Africa, Latin America and the Caribbean, and Western, Central Europe and North America. More specifically, most of the included HIV prevalence studies were published before 2010. Indeed, only two studies have been conducted in the last five years on HIV prevalence among LDTDs [46,73]. This is an indication of a decelerating or stagnated body of evidence on HIV among LDTDs in different parts of the world. Such a scenario is likely to mask and drive a silent disproportionately high burden of HIV in this population. Thus, more studies and surveillance are needed in individual countries to generate reliable up-to-date HIV prevalence estimates for more appropriate targeted interventions.

Several limitations should be considered when interpreting the findings of this review. One, the review included all existing studies on HIV prevalence among LDTDs since the start of the epindemic. Thus, a portion of the data may not be up-to-date. However, given the lack of any existing review of this kind, the current study was warranted. Second, the review focused on studies only published in the English language, meaning that some data published in other languages may have been left out, thus possibly underestimating or overestimating the global burden of HIV among LDTDs. The high heterogeneity in the included studies may limit the generalizability of the findings especially from regions where there was a huge scarcity of data. The diversity of study settings, year of publication, sample size, and socioeconomic factors among the study sample may also limit the generalizability of the findings. Lastly, some databases that are not open access were not searched for the information included in the current study. It may be that some important information in those databases was left out in this review.

## Conclusion

The global burden of HIV among LDTDs is 3.82%. This burden is highest in sub-Saharan Africa at 14.34%, followed by Asia and the Pacific at 2.14%, and the lowest in Western, Central Europe and North America at 0.17%. More LDTD-centred research and surveillance on HIV prevalence is recommended in individual countries. This would be pivotal in instituting tailored national and regional strategies, policies and interventions suitable for preventing new HIV infections among LDTDs.

## Data Availability

All the data used in developing this manuscript are available upon request from the corresponding author.

## Acknowledgements

We acknowledge the faculty of Nursing, Jomo Kenyatta University of Agriculture and Technology for their input during the preliminary presentation of this study.

## Authors’ contributions

**Conceptualization:** Cyrus Mutie, Berrick Otieno, Grace Mbuthia

**Design and Methodology**: Cyrus Mutie, Berrick otieno, Grace Mbuthia

**Formal analysis**: Cyrus Mutie, Berrick Otieno

**Supervision:** Elijah Mwangi, Rosemary Kawira, Albanus Mutisya, John Gachohi, Grace Mbuthia

**Writing – original draft:** Cyrus Mutie

**Writing – review and editing**: Elijah Mwangi, Rosemary Kawira, Albanus Mutisya, John Gachohi, Grace Mbuthia

## Supporting Information

**S1 Table. A PRISMA checklist showing the specific guideline requirements and page of location in the manuscript. DOCX.**

**S1 File. Search string from PubMed and PubMed Central (PMC). DOCX.**

**S2 Table. Summary of the 43 included studies on the global burden of HIV among Long-distance truck drivers. DOCX.**

**S3 Table. Forest plots showing sub-group analysis by regions, pooled prevalence and heterogeneity scores. DOCX.**

**S4 Table. Critical appraisal results for quality of the 39 included cross-sectional (prevalence) studies based on Joanna Briggs Checklist. XLSX.**

**S5 Table. Quality appraisal for the 2 included Cohort Studies based on New-ottawa Scale for assessing Quality and Bias for Cohort studies. XLSX.**

**S6 Table. Quality appraisal for the 2 included non-randomised interventional studies using Joanna Brigs Checklist for non-randomized studies. XLSX.**

## Notes

### Competing Interest Statement

The authors have declared no competing interest.

### Funding Statement

The study did not recieve any funding.

### Author Declarations

This is a systematic review and meta-analysis manuscript and did not require ethical approval.

